# Beyond Walking Speed: Estimating and Classifying Post-Stroke Gait Propulsion From Standardized Clinical Measures

**DOI:** 10.64898/2026.09.30.26364361

**Authors:** Jie Fei, Ruoxi Wang, Louis N. Awad

## Abstract

**Background:** Gait propulsion (anterior ground reaction force) is commonly impaired after stroke. Force plates and wearable sensors can quantify propulsion but are not routinely available in clinical settings. Walking speed alone does not adequately capture the heterogeneity of propulsion deficits.

**Objective:** To determine whether standardized clinical measures can estimate post-stroke propulsion metrics and classify propulsion impairment.

**Methods:** Forty individuals post-stroke participated: 33 for model development and cross-validation (11 with repeated observations for longitudinal validation) and 7 for independent-cohort validation. Clinical measures included the 6-Minute Walk Test (6MWT), comfortable walking speed (CWS), Functional Gait Assessment (FGA), Fugl-Meyer Assessment of the Lower Extremity (FMA-LE), Short Physical Performance Battery (SPPB), Timed Up and Go test (TUG), and stroke chronicity. Regression models were developed to estimate three propulsion metrics: paretic propulsion peak and impulse, and interlimb propulsion symmetry. Model performance was evaluated using R^2^ and RMSE. Propulsion impairment was classified from symmetry estimates using a prespecified cutoff of 31.5%, with classification accuracy reported.

**Results:** Models using the 6MWT, SPPB, TUG, and chronicity estimated paretic propulsion peak (R^2^ = 0.77, RMSE = 3.00 %body weight) and impulse (R^2^ = 0.81, RMSE = 0.65 %body weight·%stance phase). FMA-LE, SPPB, TUG, and chronicity estimated propulsion symmetry (R^2^ = 0.75, RMSE = 8.56%) and classified propulsion impairment with 84.8% accuracy. Classification accuracy was 91.7% in longitudinal validation and 85.7% in independent-cohort validation.

**Conclusions:** A multidomain clinical assessment framework may support practical and accurate estimation and classification of post-stroke propulsion impairment in clinical settings where specialized instrumentation is unavailable.

## 1 Introduction

Propulsion, defined as the anterior component of the ground reaction force (GRF) that advances the body forward during walking, is essential to safe and efficient locomotion.^1^ After stroke, muscle weakness, spasticity, and sensorimotor deficits substantially compromise the paretic limb’s capacity to produce propulsive force.^2^ This impairment contributes to slower and more asymmetrical gait, reduced endurance, and increased metabolic cost, limiting community ambulation and quality of life.^1–4^

Propulsion has therefore emerged as a central target of post-stroke gait rehabilitation.

Despite its importance, propulsion remains rarely quantified in clinical practice due to the absence of accessible measurement approaches.^2^ Direct measurement typically requires force plates or instrumented treadmills, which are commonly unavailable in clinical settings. Although indirect approaches using wearable motion sensors and pressure-sensing insoles have been developed to estimate propulsion,^5–7^ their adoption remains limited by cost, additional workload burden, and technical demands.^8,9^ Consequently, clinicians rely predominantly on walking speed and observed gait deviations, which require substantial training and practice, to grossly infer propulsion impairments.^2,10^ However, given the heterogeneity of post-stroke gait, individuals with comparable walking performance may exhibit markedly different underlying propulsion impairment driven by distinct compensatory strategies, increasing the risk of mischaracterization and suboptimal treatment.^1–3,11^

We posit that routinely collected, standardized clinical measures that provide complementary insights into post-stroke impairment could serve as feasible proxies for estimating propulsion, especially when considered together. Indeed, clinical tests such as the 6-Minute Walk Test (6MWT),^1,4^ 10-Meter Walk Test (10MWT),^12^ Functional Gait Assessment (FGA),^13^ Fugl-Meyer Assessment of the Lower Extremity (FMA-LE),^14^ Short Physical Performance Battery (SPPB),^15^ and Timed Up and Go test (TUG) ^16^ capture complementary domains spanning walking capacity, motor impairment, and functional mobility. Although none of these assessments directly measure propulsion, their multidimensional nature may encode underlying propulsion deficits.

Building on our prior work demonstrating that combining comfortable walking speed (CWS) with the FGA improves propulsion estimation and impairment classification compared with walking speed alone,^13^ the present study evaluated whether a broader, multidomain set of standardized clinical measures can further improve characterization of post-stroke gait propulsion impairment. Specifically, we developed estimation models incorporating standardized clinical measures, together with stroke chronicity as a contextual variable, to estimate propulsion metrics and classify propulsion impairment severity. We further evaluated reduced models to determine whether useful performance could be maintained when individual clinical measures were unavailable, and we assessed estimation and classification performance using cross-validation, longitudinal observations, and an independent validation cohort. Together, this multidomain clinical framework aims to advance a practical approach for estimating and classifying propulsion impairment when direct force-plate measurement is not available.

## 2 Methods

This retrospective estimation model development and validation study analyzed data from 40 individuals with chronic stroke, collected between March 4, 2021, and December 12, 2025. Inclusion criteria were age ≥18 years, history of stroke ≥6 months prior to enrollment, ability to walk for at least 2 minutes without physical assistance for body-weight support (assistive devices permitted), and ability to communicate with study investigators. Exclusion criteria included NIH Stroke Scale scores >1 on item 1b or >0 on item 1c, pain limiting walking, neglect or hemianopia, unexplained dizziness within the previous 6 months, serious musculoskeletal, cardiovascular, skin, pulmonary, or neurological comorbidities that could interfere with participation, and more than two falls in the previous month.^13^ The study was approved by the Boston University Charles River Campus Institutional Review Board, and all participants provided written informed consent.

### 2.1 Data collection

Data were collected in an indoor laboratory with a 26.6-meter oval track, including six floor-embedded force plates (Bertec Corp., Columbus, OH, USA) along a 10-meter straight section.^17^ Participants completed clinical assessments administered by licensed physical therapists, including the 10MWT, 6MWT, FGA, FMA-LE, SPPB, and TUG. Three 10MWT trials were performed at self-selected CWS along the straight section. One 6MWT was completed around the track. Ground reaction forces were captured at 2000 Hz in Qualisys Track Manager (2024.3; Qualisys AB, Göteborg, Sweden) each time participants crossed the force plates during the 6MWT.

### 2.2 Data processing

CWS was calculated as the mean walking speed across three 10MWT trials. GRF data from the 6MWT were low-pass filtered (second-order Butterworth, 10 Hz cutoff). Stance phases were identified using a 5% body weight (%BW) threshold, and the GRF data were segmented accordingly.^5^ Three stride-level propulsion metrics were calculated from the anterior GRF: paretic propulsion peak, paretic propulsion impulse, and propulsion symmetry. Propulsion peak (%BW) was defined as the maximum anterior GRF normalized to body weight, and propulsion impulse (%BW·%stance phase, %BW·%sp) was calculated as the integral of the body-weight-normalized positive anterior GRF over the normalized stance phase. Interlimb propulsion symmetry (%) was defined as the ratio of paretic propulsion impulse to the sum of bilateral propulsion impulses.^3,5,13^ Pooled stride-level metric data per participant exceeding three standard deviations from the mean were excluded, and the remaining values were averaged. Propulsion symmetry values approaching 50% indicate less propulsion impairment.^3^

### 2.3 Data analysis

Bivariate relationships among predictors and propulsion metrics were assessed before modeling. Normality was evaluated using the Lilliefors test (α = 0.05).^18^ Pearson or Spearman correlations were used as appropriate.^19^ Predictor multicollinearity was assessed using variance inflation factors (VIF), with VIF >10 indicating exclusion.^20^

To determine whether standardized clinical measures could estimate propulsion, stepwise linear regression was used to develop separate models for paretic propulsion peak, paretic propulsion impulse, and propulsion symmetry from seven candidate predictors: 6MWT, CWS, FGA, FMA-LE Total E-F (representing motor function; hereafter FMA-LE),^21^ SPPB, TUG, and chronicity.^13,22–24^ Model performance was evaluated using R^2^, adjusted R^2^, root mean squared error (RMSE), and model-level *p*- values. Regression coefficients with corresponding *p*-values were reported for all retained predictors. Model generalizability was assessed using 10-fold cross-validation. Within each fold, stepwise predictor selection and model fitting were performed using only the training data, and the resulting model was applied to the held-out observations. Cross-validated R^2^ (CV-R^2^) and RMSE (CV-RMSE) were reported.^24^

To determine whether estimated propulsion symmetry could classify propulsion impairment severity, estimated symmetry values were categorized as mild or severe using a prespecified cutoff of 31.5% derived from prior work (>31.5% = mild).^3,25,26^ Classification was compared with force-plate-derived impairment severity using overall accuracy, sensitivity, specificity, and confusion matrices. Confidence intervals (95% CIs) for these proportions were computed using the Wilson score interval.

To evaluate whether useful performance could be maintained when some clinical measures were unavailable, reduced propulsion symmetry models were generated using subsets of the predictors retained in the full propulsion symmetry model, with chronicity retained in all reduced models. For each reduced model, estimation performance was evaluated using CV-R^2^ and CV-RMSE, and classification performance was evaluated using overall accuracy (with 95% CIs).

Model performance for propulsion symmetry estimation and propulsion impairment classification was further evaluated in two validation settings: longitudinal validation using repeat observations from participants in the model-development cohort, and independent-cohort validation using data from separate participants.^24,27,28^ When predictors required by the full model were unavailable, the corresponding reduced model was applied. Estimation performance was quantified using R^2^ and RMSE, and classification performance was evaluated using overall accuracy (with 95% CIs) and confusion matrices. All analyses were performed in MATLAB (R2024a; MathWorks, Inc., Natick, MA, USA).

## 3 Results

A total of 40 individuals post-stroke were included. Thirty-three participants were used for model development and cross-validation (age 61.6 ± 10.4 years; chronicity 5.7 ± 3.7 years; 21 males, 12 females), of whom 11 contributed repeat observations for longitudinal validation. A separate group of 7 participants was used for independent-cohort validation (age 59.3 ± 11.7 years; chronicity 5.7 ± 3.3 years; 7 males). Detailed participant characteristics are summarized in **Supplementary Table 1**.

Correlations between predictors and propulsion metrics are summarized in **Supplementary Figure 1A**. Paretic propulsion peak and paretic propulsion impulse were highly correlated (r = 0.98), and both were strongly associated with propulsion symmetry (r = 0.82 and 0.86, respectively). Clinical measures showed varying associations with the three propulsion outcomes: paretic propulsion peak was most strongly associated with 6MWT (r = 0.75), paretic propulsion impulse with TUG (r = −0.76), and propulsion symmetry with FMA-LE (r = 0.59). No predictor exceeded the VIF threshold of 10 (**Supplementary Figure 1B**).

Stepwise regression results are summarized in **Table 1**. All models were significant (*p* < 0.001), with all retained predictors contributing significantly (*p* < 0.05). Paretic propulsion peak was estimated using 6MWT, SPPB, TUG, and chronicity (R^2^ = 0.77, RMSE = 3.00 %BW), and the same measures were retained in the paretic propulsion impulse model (R^2^ = 0.81, RMSE = 0.65 %BW·%sp). In contrast, propulsion symmetry was estimated using FMA-LE, SPPB, TUG, and chronicity (R^2^ = 0.75, RMSE = 8.56%). Cross-validated performance was consistent with model fit, and estimated versus observed values are shown in **Supplementary Figure 2**.

**Table 1.** Stepwise regression and cross-validation results for three propulsion metrics.

| Response | Model statistics |  |  |  | Predictor statistics |  |  | Cross-validation |  |
| --- | --- | --- | --- | --- | --- | --- | --- | --- | --- |
| | $R^2$ | Adj $R^2$ | RMSE | $p$ | Predictor | $\beta$ | $p$ | $R^2$ | RMSE |
| Paretic propulsion peak (%BW) | 0.77 | 0.74 | 3.00 | <0.001*** | Intercept | 20.62 | 0.003** | 0.72 | 3.05 |
|  |  |  |  |  | 6MWT | 0.04 | <0.001*** |  |  |
|  |  |  |  |  | SPPB | -1.10 | 0.006** |  |  |
|  |  |  |  |  | TUG | -0.69 | <0.001*** |  |  |
|  |  |  |  |  | Chronicity | -0.58 | <0.001*** |  |  |
| Paretic propulsion impulse (%BW·%sp) | 0.81 | 0.78 | 0.65 | <0.001*** | Intercept | 6.04 | <0.001*** | 0.76 | 0.68 |
|  |  |  |  |  | 6MWT | 0.01 | <0.001*** |  |  |
|  |  |  |  |  | SPPB | -0.29 | 0.001** |  |  |
|  |  |  |  |  | TUG | -0.19 | <0.001*** |  |  |
|  |  |  |  |  | Chronicity | -0.15 | <0.001*** |  |  |
| Propulsion symmetry (%) | 0.75 | 0.72 | 8.56 | <0.001*** | Intercept | 112.69 | <0.001*** | 0.69 | 8.78 |
|  |  |  |  |  | FMA-LE | 0.96 | 0.025* |  |  |
|  |  |  |  |  | SPPB | -5.64 | <0.001*** |  |  |
|  |  |  |  |  | TUG | -3.16 | <0.001*** |  |  |
|  |  |  |  |  | Chronicity | -1.01 | 0.025* |  |  |

\* $p < 0.05$ ; \*\* $p < 0.01$ ; \*\*\* $p < 0.001$
**Abbreviations:** $R^2$ = coefficient of determination; Adj $R^2$ = adjusted coefficient of determination; RMSE = root mean square error; $p$ = $p$ -value; $\beta$ = regression coefficient estimate.

Classification performance for propulsion impairment severity based on estimated propulsion symmetry is shown in **Figure 1**. Of the 33 participants, 28 were correctly classified (accuracy = 84.8%; 95% CI: 69.1–93.3%). Sensitivity for detecting severe impairment was 81.0% (17/21; 95% CI: 60.0–92.3%), with 4 participants misclassified as mild. Specificity for identifying mild impairment was 91.7% (11/12; 95% CI: 64.6–98.5%), with 1 participant misclassified as severe.

**Figure 1.**
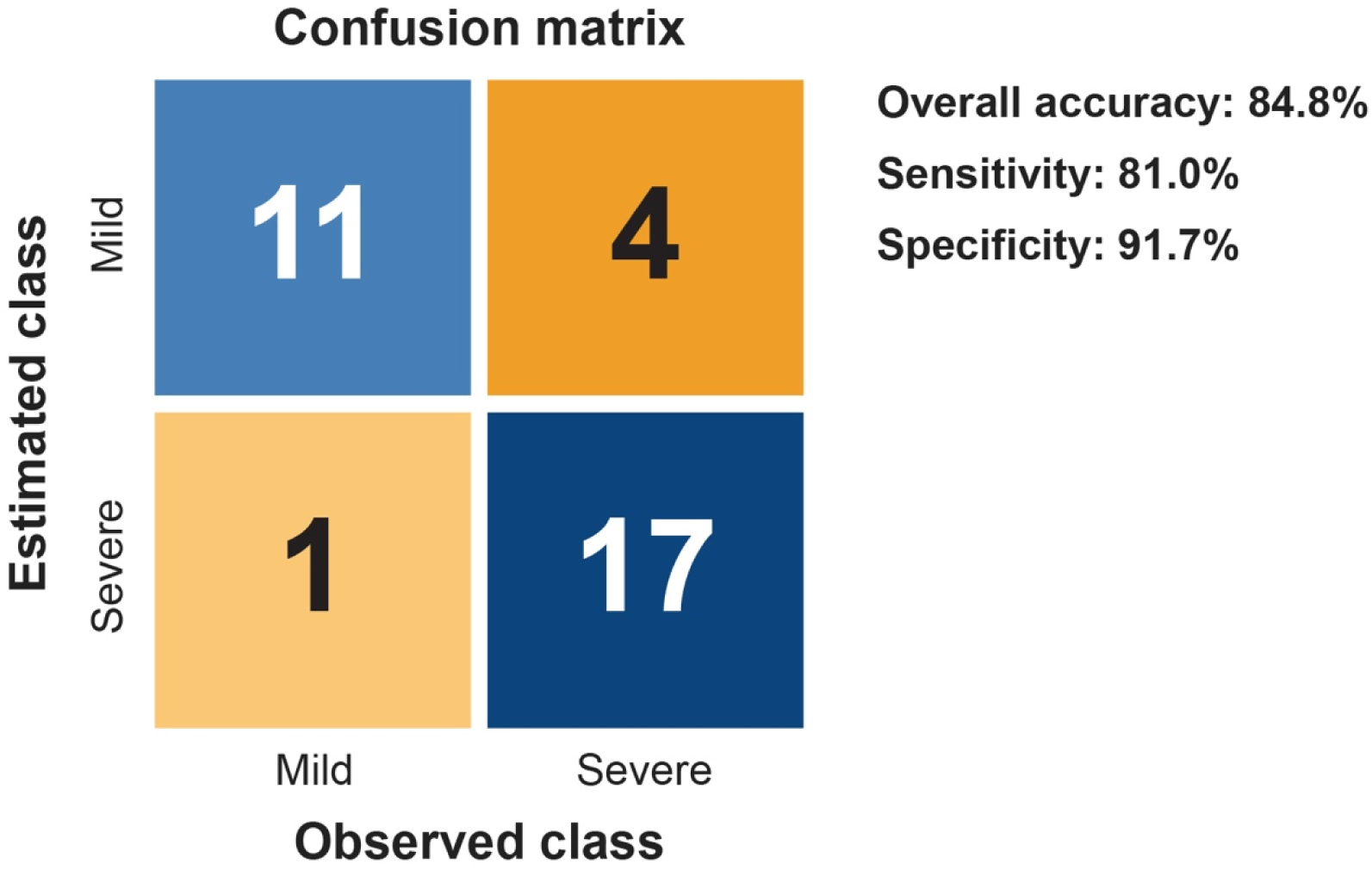
Classification performance for propulsion impairment severity. The confusion matrix shows agreement between impairment severity classified from estimated versus observed propulsion symmetry, with overall accuracy, sensitivity, and specificity reported.

Results of the predictor subset analysis, including model equations, are presented in **Supplementary Table 2**. Compared with the full stepwise regression model (CV-R^2^ = 0.69, CV-RMSE = 8.78%, accuracy = 84.8%), Reduced model 1, which excluded FMA-LE and retained SPPB, TUG, and chronicity, achieved a slightly lower CV-R^2^ (0.62) and higher CV-RMSE (9.70%) but the highest classification accuracy (87.9%; 95% CI: 72.7–95.2%). The remaining reduced models (models 2–6) showed progressively lower CV-R^2^ (0.39 to −0.08) and lower classification accuracy (78.8% to 63.6%).

Longitudinal and independent-cohort validation results for propulsion symmetry estimation and impairment classification using the Full model or Reduced model 1 are shown in **Figure 2**. In the longitudinal validation, propulsion symmetry was estimated with R^2^ = 0.65 and RMSE = 7.70%, and impairment severity was classified with 91.7% accuracy (95% CI: 64.6–98.5%). Eleven participants contributed 12 repeat visits (11 second visits and 1 third visit). The Full model was applied for 5 visits and Reduced model 1 for 7 visits with missing FMA-LE data. All mild cases (6/6) and five of six severe cases (5/6) were correctly classified. In the independent-cohort validation, propulsion symmetry was estimated with R^2^ = 0.79 and RMSE = 7.47%, and impairment severity was classified with 85.7% accuracy (95% CI: 48.7–97.4%). Reduced model 1 was used for all seven participants because FMA-LE data were unavailable. All mild cases (3/3) and three of four severe cases (3/4) were correctly classified.

**Figure 2.**
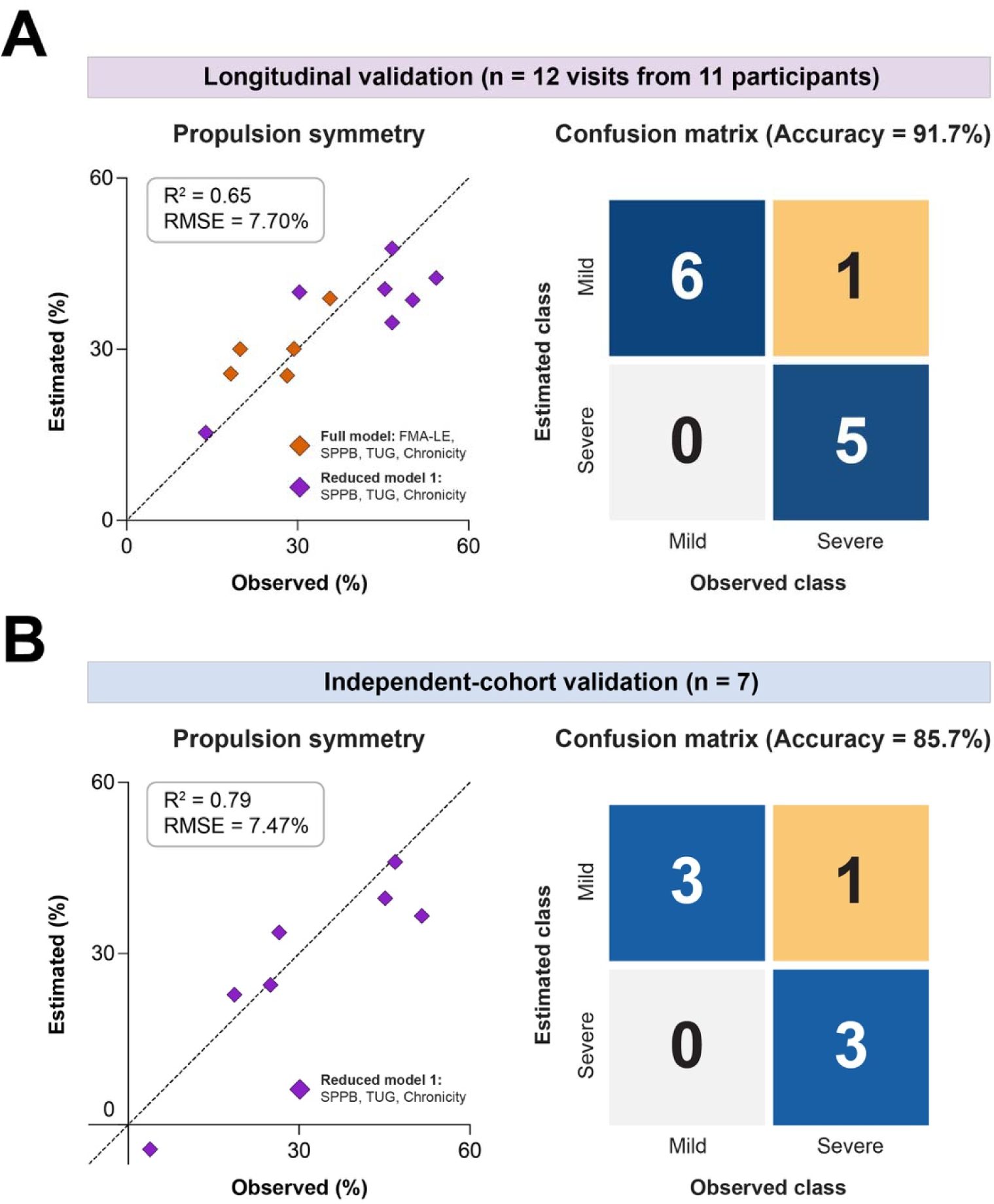
Model performance for propulsion symmetry estimation and propulsion impairment classification in longitudinal and independent-cohort validation. Estimated versus observed propulsion symmetry values (left) and confusion matrices (right) are shown for **(A)** longitudinal validation and **(B)** independent-cohort validation. Propulsion symmetry estimates were derived using the Full model (orange diamonds) or Reduced model 1 (purple diamonds). Reduced model 1 was applied when FMA-LE data were unavailable. The dashed line indicates the line of identity.

Longitudinal trajectories of observed and estimated propulsion symmetry, with corresponding propulsion impairment severity classifications across the model development and longitudinal validation visits of the 11 longitudinal participants, are shown in **Figure 3**. Estimated classifications were consistent with force-plate-derived classifications across all visits for participants L1–L4 and L7–L11, whereas participants L5 and L6 were misclassified at the first visit only.

**Figure 3.**
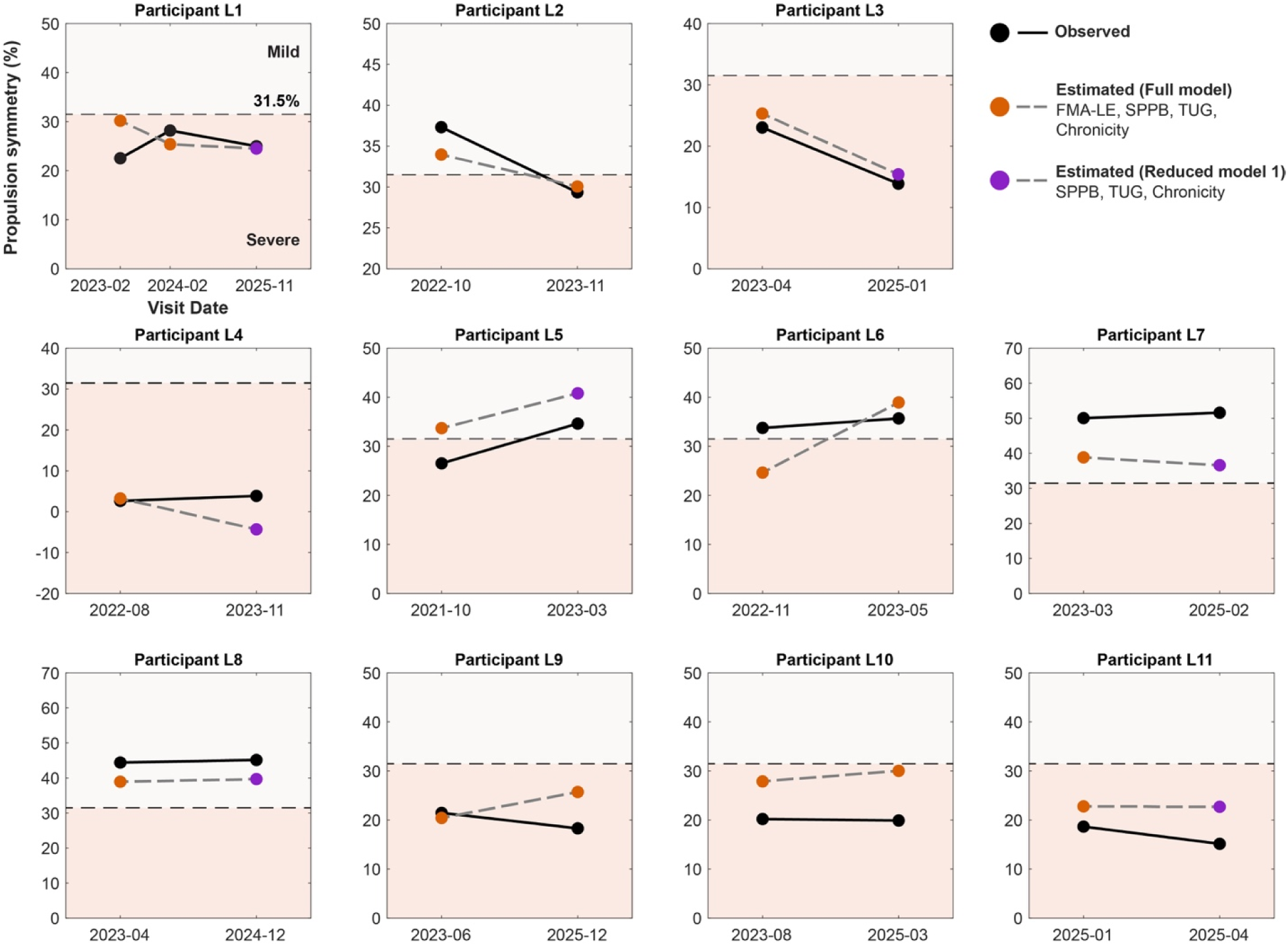
Longitudinal tracking of observed and estimated propulsion symmetry with corresponding impairment classification across visits. Each panel shows observed values (black) and estimates from the Full model (orange) or Reduced model 1 (purple) across two or three chronologically ordered visits for 11 participants (L1–L11). The horizontal dashed line indicates the prespecified 31.5% cutoff separating mild (>31.5%; upper gray shading) and severe (≤31.5%; lower red shading) propulsion impairment. First visits were used for model development, whereas subsequent visits were used for longitudinal validation. Reduced model 1 was applied when FMA-LE data were unavailable.

## 4 Discussion

This study extends our prior demonstration that post-stroke propulsion impairment can be characterized using standardized clinical measures beyond walking speed by showing that measures spanning walking capacity, motor impairment, and functional mobility can be integrated to estimate multiple dimensions of gait propulsion and support impairment classification when force-plate measurements are unavailable. Relative to our prior model using CWS and FGA,^13^ the current approach improved estimation across all three propulsion metrics, with greater explained variance and lower estimation error. Improvement was particularly evident for propulsion symmetry, a metric sensitive to interlimb neuromechanical differences.^1,29^ Together, these findings support the premise that a broader set of clinical measures provides complementary information about the underlying propulsion phenotype. The magnitude of estimation error also supports the potential clinical utility of this approach: the cross-validated RMSE for paretic propulsion impulse (0.68 %BW·%sp) was below the reported overground minimal detectable change of 0.90 %BW·%sp,^30^ suggesting that model error is comparable to the measurement variability inherent in gold-standard instrumented analysis.

To extend the utility of these estimates from continuous propulsion characterization to clinically interpretable impairment classification, we applied a prespecified cutoff, derived from the Brunnstrom stages of stroke recovery,^31^ to classify propulsion impairment severity.^25^ We achieved an overall classification accuracy of 84.8%, exceeding the 72.5% accuracy of our prior CWS- and FGA-based approach, which relied on a less generalizable in-sample threshold derived from paretic propulsion peak.^13^ Longitudinal and independent-cohort validation accuracies of 91.7% and 85.7%, respectively, further support the robustness of classification performance across time points and new participants with varying propulsive capacities.^27,28^ Recognizing that the complete predictor set may not always be available in routine clinical practice, we also evaluated reduced models with predictor subsets. A reduced model comprising SPPB, TUG, and chronicity maintained competitive performance, providing a practical fallback when clinical data availability is constrained.

Despite starting from seven candidate predictors, the final stepwise models retained only four predictors for each propulsion metric, with CWS and FGA excluded from all models. The exclusion of CWS is consistent with accumulating evidence that walking speed and propulsion represent related but partially dissociable constructs.^3,12,13,32^ The FGA, although informative in prior work, was superseded by the TUG and SPPB, which appear to capture dynamic balance and functional mobility with greater specificity to propulsion-related demands.^15,33,34^ The TUG was among the most consistently retained predictors across models and demonstrated a strong negative association with propulsion metrics, which may reflect its integration of multiple mobility subtasks that require coordinated bilateral propulsive force generation.^33,35,36^ The SPPB was also retained in all models. Despite its weak bivariate association with propulsion symmetry, the SPPB assesses lower-extremity strength, balance, and habitual gait speed—domains that collectively index neuromuscular capacity and coordination underlying bilateral propulsive capacity and likely capture residual variance in propulsion symmetry not explained by the other retained predictors.^34,37^

The 6MWT emerged as the primary predictor for paretic propulsion peak and impulse, consistent with its established role as a measure of walking endurance and sustained propulsion output.^4,38,39^ Notably, the 6MWT was replaced by the FMA-LE in the propulsion symmetry model, which may suggest that propulsion symmetry depends more directly and strongly on corticospinal control of distal plantarflexor function captured by the FMA-LE than on global walking endurance.^1,14,32,40^ Supporting this interpretation, the six individuals with observed severe propulsion impairment who were misclassified as mild across the development and validation analyses had an average 6MWT distance greater than the model-development cohort mean (368.94 vs. 319.20 m). One possible explanation is that 6MWT performance can be sustained through compensatory strategies despite persistent propulsive asymmetry, thereby biasing impairment classification toward less severe categories in individuals who maintain endurance through imbalanced propulsive mechanics.^1,2,32,41^ Chronicity was also retained in all models, with negative regression coefficients, despite negligible bivariate correlations (r ≤ 0.06), a pattern consistent with a statistical suppressor effect.^42^ Its negative partial association likely reflects progressive consolidation of compensatory strategies over time, which attenuates propulsive output independently of variance shared with other predictors.^2,43,44^

Finally, to support practical application of these findings, an interactive spreadsheet implementing the validated Full model and Reduced model 1 for propulsion symmetry estimation and propulsion impairment classification is provided, with instructions and a link, in **Supplementary Figure 3**.

### 4.1 Limitations and future directions

The sample comprised exclusively individuals with chronic stroke, limiting generalizability to subacute or acute stroke populations, in whom spontaneous neurological recovery may produce different and more rapidly evolving relationships between clinical measures and propulsion.^45,46^ Because the linear regression models are unconstrained, estimated propulsion symmetry may occasionally fall outside the physiologically plausible range (e.g., below 0%), particularly for individuals with predictor profiles near or beyond the range represented in the development sample. Although complementary validation frameworks were employed, statistical power remained limited by the small sample size. This is reflected in the wide 95% CIs around classification accuracy, particularly in the independent cohort (48.7–97.4%), indicating that validation-cohort results should be interpreted as preliminary evidence of generalizability rather than precise performance estimates. Furthermore, all clinical measures were administered by physical therapists within a single research laboratory. Because several of these measures involve rater judgment, scores obtained in other settings or by other raters may differ, which may affect the generalizability of this approach. Prospective, multi-site evaluation in larger, more demographically and clinically diverse cohorts is therefore needed to establish clinical validity at scale.^44^ Additionally, it remains to be determined whether changes in clinical measure scores following propulsion-targeted interventions translate into changes in estimated propulsion that are sufficiently sensitive and accurate to support monitoring of propulsion impairment over the course of treatment.^2,47^

These estimation models should not be interpreted as substitutes for instrumented biomechanical assessment when direct measurement is available. Rather, they provide a lower-burden approach for clinically characterizing propulsion in settings where force plates or instrumented treadmills are not accessible.^9,48^ Wearable sensing approaches may provide an intermediate option when more direct or repeated biomechanical measurement is needed.^5,6^ Future work should also determine whether incorporating additional clinical measures, leveraging subcomponents of measures, or integrating a minimal sensor configuration can narrow the remaining accuracy gap while preserving clinical implementation feasibility.^49,50^

## 5 Conclusions

Standardized clinical measures spanning walking capacity, motor impairment, and functional mobility can be integrated to estimate post-stroke gait propulsion and support classification of propulsion impairment severity. These findings extend prior work demonstrating that propulsion can be characterized beyond walking speed alone and support a multidomain clinical assessment framework when direct force-plate measurement is unavailable. Longitudinal and independent-cohort validation further provide preliminary evidence that model performance extends beyond the development sample.

## Data Availability

All data produced in the present study are available upon reasonable request to the authors

## Acknowledgments

The authors thank the study participants and the members of the Neuromotor Recovery Laboratory at Boston University for their assistance with data collection.

## Author contributions

Jie Fei: Conceptualization; data curation; formal analysis; funding acquisition; investigation; methodology; project administration; software; validation; visualization; writing – original draft; writing – review & editing. Ruoxi Wang: Formal analysis; methodology; writing – review & editing. Louis N. Awad: Conceptualization; funding acquisition; methodology; project administration; resources; supervision; writing – review & editing.

## Statements and declarations

### Ethical considerations

This study was conducted in accordance with the Declaration of Helsinki and approved by the Boston University Charles River Campus Institutional Review Board (protocol number: #4440; approval date: April 10, 2016).

### Consent to participate

Written informed consent was obtained from all participants involved in the study.

### Consent for publication

Not applicable.

### Data availability statement

The data supporting the findings of this study are available from the corresponding author upon reasonable request.

### Declaration of generative AI and AI-assisted technologies in the writing process

During the preparation of this work, the authors used ChatGPT in order to improve grammar, clarity, and language style. After using this tool, the authors reviewed and edited the content as needed and take full responsibility for the content of the publication.

