## Supplementary for "Beyond Walking Speed: Estimating and Classifying Post-Stroke Gait Propulsion From Standardized Clinical Measures"

***Supplementary material***

**Supplementary Table 1.** Participant demographic and clinical characteristics.

| Characteristic | Model development,  cross-validation, and  longitudinal validation (n = 33) | Independent-cohort  validation (n = 7) |
| --- | --- | --- |
| **Demographic**  Age, y, mean (SD; range)  BMI, kg/m^2^, mean (SD; range)  Chronicity, y, mean (SD; range)  Paretic leg (L/R), n  Sex (M/F), n  **Clinical**  6MWT, m, mean (SD; range)  CWS, m/s, mean (SD; range)  FGA, mean (SD; range)  FMA-LE, mean (SD; range)  SPPB, mean (SD; range)  TUG, s, mean (SD; range) | 61.64 (10.36; 41.00–83.00)  28.89 (5.83; 19.18–47.30)  5.70 (3.74; 0.90–15.50)  16/17  21/12  319.20 (103.93; 146.63–532.94)  0.89 (0.28; 0.45–1.45)  16.67 (6.05; 6.00–30.00)  23.00 (5.94; 9.00–33.00)  9.06 (2.33; 3.00–12.00)  14.22 (5.58; 7.37–30.00) | 59.29 (11.66; 43.00–76.00)  30.58 (4.31; 24.96–38.60)  5.67 (3.26; 1.17–10.90)  6/1  7/0  331.10 (155.18; 134.56–614.74)  0.65 (0.20; 0.47–1.03)  15.25 (5.97; 11.00–24.00)*  —**^†^**  8.29 (3.25; 3.00–12.00)  13.00 (4.56; 8.92–21.96) |

**Abbreviations:** y = years; BMI = body mass index; kg/m^2^ = kilograms per square meter; L = left; R = right; M = male; F = female; 6MWT = 6-Minute Walk Test; m = meters; CWS = comfortable walking speed; m/s = meters per second; FGA = Functional Gait Assessment; FMA-LE = Fugl-Meyer Assessment of the Lower Extremity (motor function); SPPB = Short Physical Performance Battery; TUG = Timed Up and Go test; s = seconds.

* FGA data were unavailable for 3 participants.

† FMA-LE data were unavailable for all 7 participants.

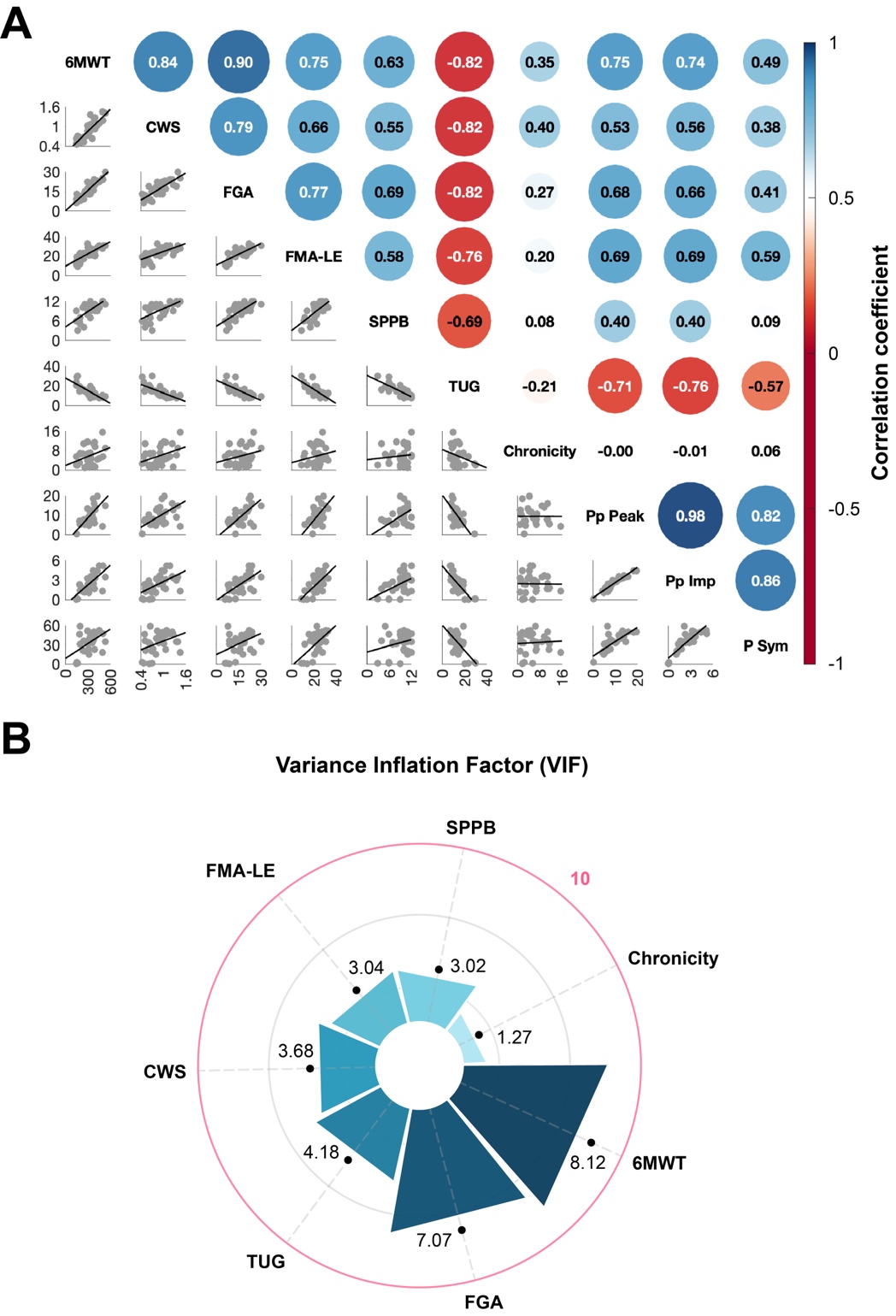

**Supplementary Figure 1.** Pairwise correlations among clinical measures and propulsion metrics, and assessment of predictor multicollinearity. **(A)** Correlation matrix showing Pearson or Spearman correlation coefficients (selected based on normality) between all variables: 6-Minute Walk Test (6MWT, m), comfortable walking speed (CWS, m/s), Functional Gait Assessment (FGA), Fugl-Meyer Assessment of the Lower Extremity motor function (FMA-LE), Short Physical Performance Battery (SPPB), Timed Up and Go test (TUG, s), chronicity (years), paretic propulsion peak (Pp Peak, %body weight), paretic propulsion impulse (Pp Imp, %body weight·%stance phase), and propulsion symmetry (P Sym, %). The upper triangle displays correlation coefficients, with circle size and color indicating the magnitude and direction of each association (blue, positive; red, negative; range −1 to 1). The lower triangle shows pairwise scatterplots with fitted regression lines. **(B)** Radar chart of variance inflation factors (VIF) for all candidate predictors. The outer pink circle indicates the threshold for severe multicollinearity (VIF = 10).

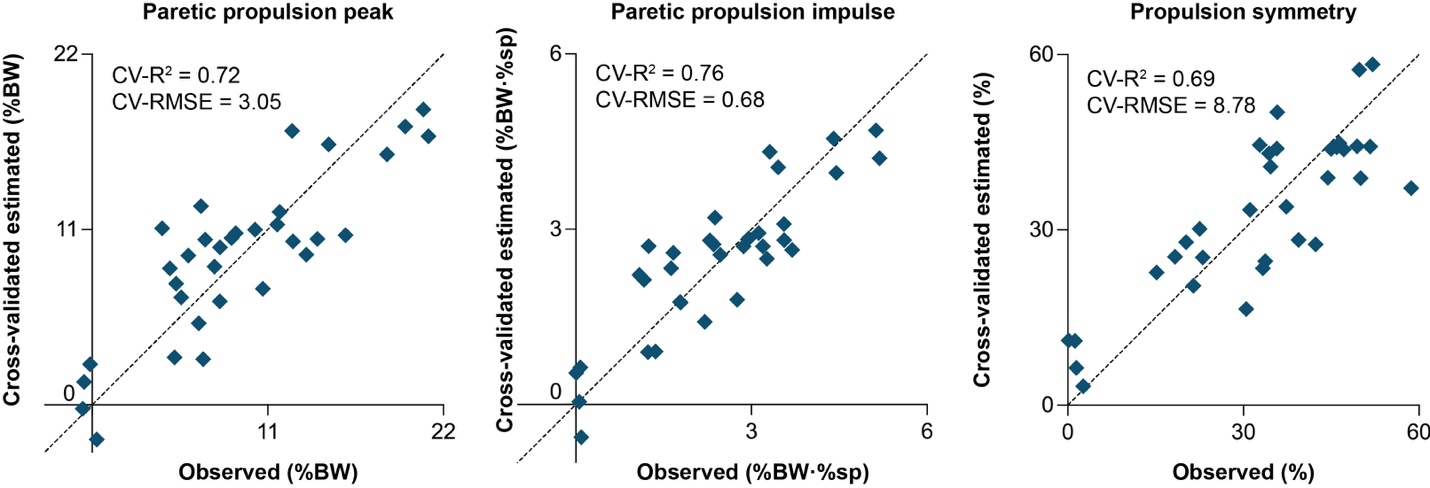

**Supplementary Figure 2.** Cross-validated estimated versus observed values for the three propulsion metrics. Scatter plots show estimated values from 10-fold cross-validation against observed values for paretic propulsion peak (left), paretic propulsion impulse (middle), and propulsion symmetry (right). Cross-validated performance metrics (CV-R^2^ and CV-RMSE) are reported in each panel. The dashed line indicates the line of identity. BW = body weight; sp = stance phase; CV = cross-validated.

**Supplementary Table 2.** Predictor subset analysis results for propulsion symmetry estimation and propulsion impairment classification.

| Model and regression equation | Cross-validation | | Classification accuracy (%) |
| --- | --- | --- | --- |
|  | *CV-R^2^* | *CV-RMSE (%)* |  |
| **Full model:** FMA-LE, SPPB, TUG, Chronicity  Propulsion symmetry = 112.69 + 0.957 × FMA-LE − 5.639 × SPPB − 3.157 × TUG − 1.013 × Chronicity | 0.69 | 8.78 | 84.8 |
| **Reduced model 1:** SPPB, TUG, Chronicity  Propulsion symmetry = 137.71 − 5.055 × SPPB −  3.744 × TUG − 1.006 × Chronicity | 0.62 | 9.70 | 87.9 |
| **Reduced model 2:** TUG, Chronicity  Propulsion symmetry = 66.03 − 2.072 × TUG −  0.635 × Chronicity | 0.39 | 12.35 | 78.8 |
| **Reduced model 3:** FMA-LE, TUG, Chronicity  Propulsion symmetry = 50.74 − 0.439 × FMA-LE −  1.714 × TUG − 0.618 × Chronicity | 0.36 | 12.63 | 72.7 |
| **Reduced model 4:** FMA-LE, SPPB, Chronicity  Propulsion symmetry = 3.69 + 2.202 × FMA-LE −  2.181 × SPPB − 0.285 × Chronicity | 0.26 | 13.56 | 69.7 |
| **Reduced model 5:** FMA-LE, Chronicity  Propulsion symmetry = −2.83 + 1.618 × FMA-LE −  0.252 × Chronicity | 0.23 | 13.89 | 66.7 |
| **Reduced model 6:** SPPB, Chronicity  Propulsion symmetry = 18.14 + 1.544 × SPPB +  0.145 × Chronicity | −0.08 | 16.41 | 63.6 |

**Abbreviations:** CV = cross-validated; R^2^ = coefficient of determination; RMSE = root mean square error; FMA-LE = Fugl-Meyer Assessment of the Lower Extremity (motor function); SPPB = Short Physical Performance Battery; TUG = Timed Up and Go test.

**Note:** Models are ordered by descending CV-R^2^. The Full model and Reduced model 1 are highlighted with green shading to indicate the two best-performing models across both propulsion symmetry estimation and propulsion impairment classification.

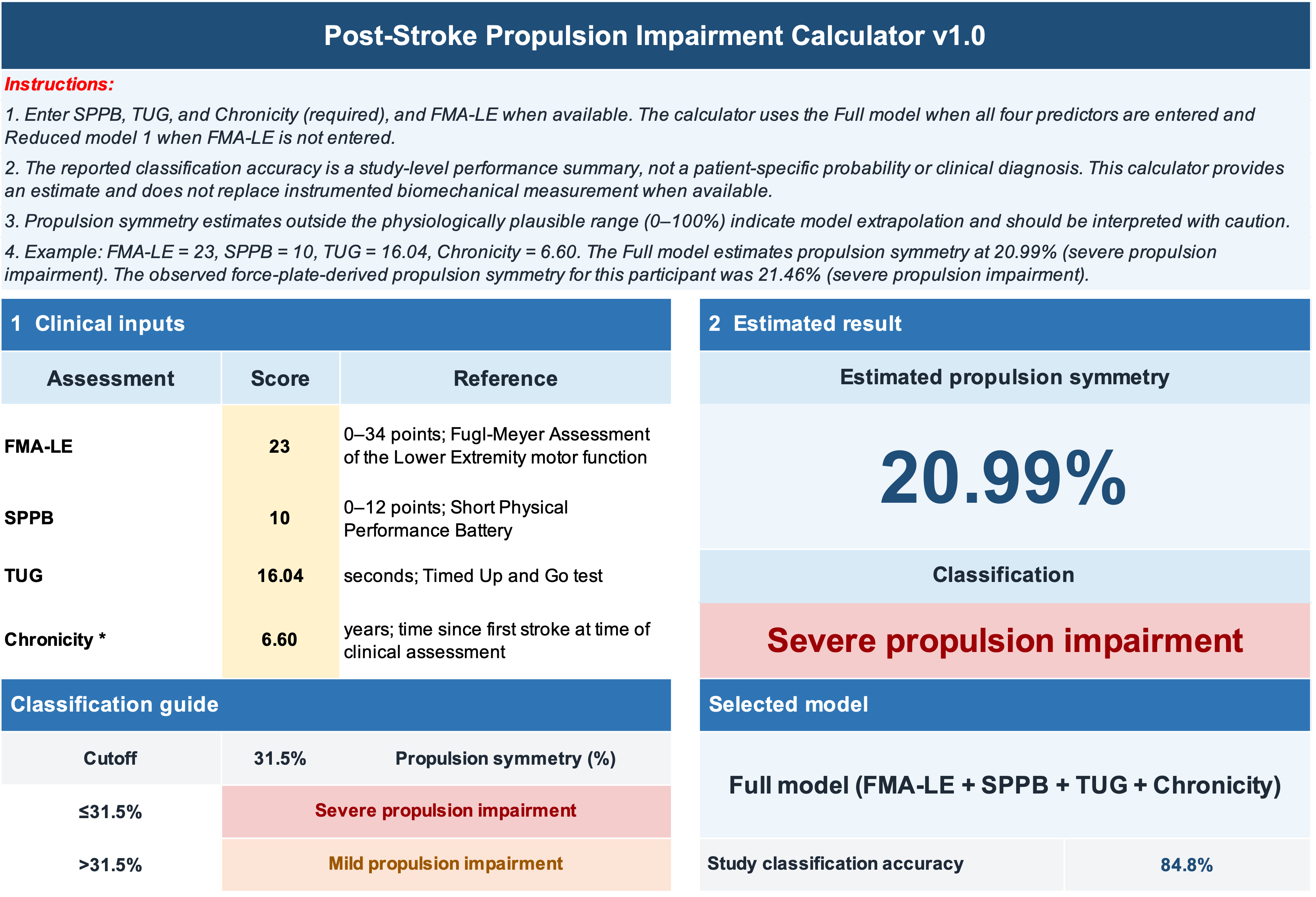

**Supplementary Figure 3.** Interactive spreadsheet for propulsion symmetry estimation and propulsion impairment classification. Users enter clinical measures including FMA-LE, SPPB, TUG, and chronicity. The calculator applies the Full model when all four predictors are available or Reduced model 1 when FMA-LE is unavailable and reports the estimated propulsion symmetry, impairment classification (mild, >31.5%; severe, ≤31.5%), selected model, and its study-level classification accuracy. Available at <https://github.com/NeuromotorRecoveryLab/Estimating-and-Classifying-Post-Stroke-Gait-Propulsion-From-Standardized-Clinical-Measures.git>.
